# Antihypertensive medication adherence and its associated factors among adult hypertensive patients at public hospitals in Addis Ababa, Ethiopia

**DOI:** 10.1101/2025.06.26.25330385

**Authors:** Habtamu Mulu Mada, Ayalnesh Zemene Yalew, Habtamu Ayalew

**Author notes:** Corresponding author: Habtamu Ayalew Mekonnen Tell; 1271.

## Abstract

**Back Ground:** Th**e** prevalence of hypertension among adults in Ethiopia is about 20.6%, and antihypertensive medication adherence is one of the indicators of treatment success for hypertensive patients. However, little is known about the current prevalence and the factors associated with antihypertensive medication adherence in the study area.

**Patients and Methods:** An institution-based cross-sectional study was conducted from March 6 to April 5, 2023 (G.C.). The sample size was 414. Four public hospitals in Addis Ababa were selected using a lottery method. To determine the number of study subjects in each hospital, a proportional allocation was used based on the total number of hypertensive patients. Respondents were selected through simple random sampling.

**Results:** The study revealed an overall adherence rate of 66.2%, with a mean age of 56.31 ± 14.61. Several factors were found to be significantly associated with adherence. These include knowledge about hypertension and its management (AOR=2.477, 95% CI: 1.499-4.093), patient-to-healthcare provider communication (AOR=2.165, 95% CI: 1.331-3.520), availability of anti-hypertension drugs in hospitals (AOR=1.778, 95% CI: 1.096-2.885), and controlled blood pressure (AOR=2.773, 95% CI: 1.707-4.507). Notably, the practice of self-blood pressure monitoring was not found to be statistically significantly associated with adherence. These findings suggest that factors such as knowledge, communication with healthcare providers, drug availability, and blood pressure control play significant roles in patient adherence to hypertension management.

**Conclusion and Recommendation:** The study highlights suboptimal medication adherence among hypertensive patients. Improving patient-to-healthcare provider communication is crucial for enhancing adherence. Educating patients about hypertension and ensuring the availability of anti-hypertension drugs in hospitals is also essential. This multifaceted approach can lead to better adherence and improved health outcomes for hypertensive patients.

## Introduction

### Background

Hypertension is systolic blood pressure equal to or above 140 mm Hg, and/or diastolic blood pressure equal to or above 90 mm Hg(1). There are numerous risk factors for hypertension, such as advanced age (>65 years), sex (male >female), heart rate (>80 beats per minute), increased body weight, diabetes, high low density lipoprotein, family history of CVD, family history of hypertension, early onset of menopause, smoking habits, psychosocial factors, and socioeconomic factors(1).

Untreated hypertension or who are treated but still have uncontrolled blood pressure are more likely to have heart and cerebrovascular disease overall(2). Because it can causes: heart disease, cerebrovascular disease, and stroke are all greatly increased by hypertension(3,4) as well as heart failure, peripheral vascular disease, renal impairment, retinal hemorrhage, and visual impairment are further serious effects of high blood pressure(1).

The prevalence of hypertension is increasing from time to time, and it is higher in low and medium-income countries than in developed countries(5). For example in Pakistan, 26.3%(6), in Uganda, 20-27%(7), and 20.6% in Ethiopia(8).

A well-known factor causing poor blood pressure management in hypertension and associated complications is failure to take drugs as directed, and continue therapy for a long period of time(9) which can lead to poor clinical outcomes(10). According to a recent study conducted in the USA, the yearly cost of drug-related morbidity and mortality as a result of non-adherence to treatment is projected to be $528.4 billion(11). And it is becoming major public as well as health care burden in low to medium income countries(5). So, in order to reduce hypertension related complications adherence, the process by which patients take and implement their medications as recommended by their health care provider(12) need to be achieved.

Due to the influence of several factors, medication adherence in people with HTN varies greatly. The development and improvement of strategies for improving individuals’ adherence, which eventually results in improved HTN control, depends on an understanding of factors determining medication adherence(13). As of 2017, research done in the USA found that patients who had good communication with their health care providers adhered three times more to their ant hypertension treatment(14). Especially those involving older individuals with HTN, which indicated that better patient-health care provider communication was linked to higher quality care and medication adherence(13,15).

According to an Italian study reported in 2019, the practice of self-blood pressure monitoring (SBPM) is related to increased adherence to medications for hypertension along with an understanding of hypertensive management(16). In the same way, in 2019, a Malaysian study reported that individuals with hypertension who self-monitor their blood pressure considerably improve their medication adherence(17).

Even though patient-to-health care providers’ communication and self-blood pressure monitoring among hypertensive patients is an important significant factor for the management and medication adherence of hypertension it is unknown in Ethiopia.

### General Objective

The aim of this study is to determine the prevalence of antihypertensive medication adherence in adult hypertensive patients at public hospitals in Addis Ababa and to identify factors associated with antihypertensive medication adherence at public hospitals in Addis Ababa, Ethiopia, 2023 GC.

### Specific Objective

✓ To Determine prevalence of antihypertensive medication adherence in adult hypertensive patients in Addis Ababa; Ethiopia,2023
✓ To identify factors associated with antihypertensive medication adherence at public hospitals in Addis Ababa; 2023

Factor like patient-to-health care provider communications and self-monitoring of blood pressure are an important aspect for the management and medication adherence of hypertension, which many studies have shown to be significant, is unknown in the study area, so the findings of this study contribute to understanding adherence to anti-hypertension medication and its associated factors in Addis Ababa.

## Methods and Materials

### Study Setting and Period, Study Design

The study took place in Addis Ababa, the capital city of Ethiopia and the location of the African Union and Economic Commission for Africa. Addis Ababa has a population of 4.8 million people (as of 2017) and is spread across an area of 540 square kilometers(18). There are 12 public hospitals that give chronic follow up services in Addis Ababa city among these Tikur Anbesa Specialized Hospital (TASH), St. Paul hospital millennium medical college (SPHMMC), St. Peter specialized hospital (SPSH) and Ras Desta Damtew memorial hospital (RDDMH) were the four public hospitals selected by lottery methods where the study carried out. An Institutional based cross-sectional study was conducted from March, o6 to April 05, 2023 in Addis Ababa Ethiopia.

#### Source population

All hypertensive patients on follow-up at public hospitals in Addis Ababa.

#### Study Population

All adult hypertensive patients on anti-hypertensive medication found at selected public hospitals during data collection and those fulfilling the inclusion criteria.

#### Study Design

Institutional based cross sectional study design was conducted from March 06 to April 05/2023.

#### Inclusion criteria

All hypertensive patients equal to or above 18 years old and who had started antihypertensive medication at least for the last six months.

#### Exclusion criteria

Seriously ill Patients, psychotic and/or unable to communicate with data collector due to other underlying medical disorder were excluded

### Sample size determination

The sample size is calculated based on objective of the study using a single population proportion formula as follows

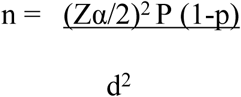

Where: n = the minimum sample size required

Zα\2 = the critical values at 95% confidence level of certainty = 1.96

d= margin error = 5

P=using the estimated proportion of good adherence to prescribed antihypertensive medications of public hospitals in North Showa Zone, North Ethiopia in 2021(4). Indicated that 56.9% of the participants were adherent.

Therefore, n = (1.96)2 ×0.569(1-0.569)/ (0.05)2

n = (1.96)2× (0.569 x 0.431) **/** (0.05)2

n=368

And by adding 10% for non-response rate, the total sample size became 414

### Sampling technique and procedure

Four hospitals were chosen by a simple random sampling method from among the entire twelve public hospitals in Addis Ababa, and 414 participants were proportionally allocated to each hospital. After assigning each respondent with the lottery method from the lists of hospital patients’ registration (a list of patients kept by the hospital that are registered at the time of diagnosis at first contact) during the vital sign check-up, the interview was taken randomly in the waiting area. To minimize recycling, participants’ cards were checked and coded.

Based on proportional allocation to size 414 study subjects was distributed to each hospital using the following formula.

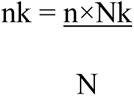

Where;

nk =required sample size from each hospital

n=the total sample size=414

Nk= number of adult hypertensive patients in each selected hospital

### Study Variables

#### Dependent: Ant-hypertensive medications adherence(adhered, not adhered)

In which the individual’s antihypertensive medication status was assessed, and it contains eight closed-ended questions, which were eight items in Morisky’s Medication Adherence Scale (MMAS-8), so the seven "yes" and "no" questions and the last five-point Likert response was used with the options "never," "once," "sometimes," "usually," and "always."(10).

#### Independent variables

**Socio-demographic related factors;** age, gender, residence (Urban and rural), marital status, educational status, and income per month

**Patient related factors;** knowledge of hypertension and its management, practice of self-blood pressure monitoring, comorbidity status, current blood pressure status, duration of treatment, and number of drugs (anti-hypertensive oral pills taken on a regular basis).

**Organizational related factors;** Patient-to-health care providers communication, health insurance usage, and drug availability in the hospitals.

### Operational Definition

#### Adherence

According to a recent researches done with Morisky Medication Adherence Scale (MMAS-8) in different country a score greater than or equal to six is considered as adherent to medication(10).

#### Non-adherence

Respondents who score less than six is considered as non-adherent to medication(10).

#### Knowledge

Participants knowledge about hypertension and its treatment which will be assessed with a scale of 12 and participants who score above the median on twelve items are considered to be knowledgeable, while scores below the median will be non-knowledgeable(19).

#### Controlled hypertension

can be defined as having a systolic blood pressure of <140 and/or a diastolic blood pressure of <90 in those on anti-hypertensive medication(20).

#### Number of pills

all types of oral ant hypertension medications classified as >2 and <2, taken on a regular basis and prescribed by health care providers(21).

#### Patient-health care provider communication

Communication between the patient and health service provider regarding patients’ clinical condition which classified as; Respondent who score equal to and above the mean for the 5 liker question of patient provider communication was considered as good patient provider communication and respondent with score below the mean for the 5 likert question of patient provider communication was considered as poor patient provider communication(22).

### Data Collection tools

Structured questionnaire and patient care review check list was used to collect data which adapted from different relevant literatures(4,7,21). The questionnaire has four parts: p-I; Socio-demographic related factors, P-II; Patient related factors, P-III: Organizational related factors and P-IV: was questions important to assess hypertensive medication adherence which was eight items Morisky Medication Adherence Scale (MMAS-8) and these were prepared in English then translated to local language Amharic and back-translated into English to maintain the consistency of the questionnaire by local language translator expertise.

### Data Collection procedures

Data collection was started after the calculated sample sizes had been allocated proportionally to each hospital. The data was collected by four BSc nurses and supervised by one another BSc nurse who had previous experience in data collection. Data collectors and supervisor were trained for two days about the consent form, how to review and collect the data. Patients who were present during data collection and eligible were interviewed by using simple random sampling technique until the calculated sample achieved while sitting in the waiting area and the remained clinical part was taken from patients check list review after examination then the filled questionnaires was checked for its completeness.

### Data Quality Assurance

The questionnaire was prepared in English and translate into Amharic language and then back to English by local language expertise. Also training was given to data collectors and supervisor for two days on how to approach study subjects and how to use the questionnaire. The principal investigator and supervisors supervised. After assessment of data collectors’ well understanding of the questionnaire the pre-test conducted on 21 participants at Yekatit 12 hospital before the actual data collection. And all of patients had understood and responded for the interview while some modification was made

### Data Processing and Analysis

Data was checked for its completeness and missing values, cleaned, cross-checked, coded, and then entered into Epi-data 4.6 Soft-ware, and exported to SPSS version 26 for statistical analysis. Statistical analysis was made at the 95% confidence level and with a 5% margin of error. The data was summarized and described by using descriptive statistics; Frequency, means, medians and standard deviation.

Bivariate and multivariable logistic regressions were used to identify associated factors with adherence status to anti-hypertensive medications. Bivariate analysis was carried out for each independent variable to identify the presence of an association with the response variable at a P-value < 0.25. Then, predictor variables that were significantly associated with response variables with a p-value of <0.25 were entered into a multivariate logistic regression model after multi-collinearity was checked by using the variance inflation factor, and the fitness of the model was also checked by the Hosmer-Lemeshow goodness of fit test. Variables with a P-value <0.05 in the multivariable analysis were considered statistically significant. And the predictor variables were identified by looking adjusted odds ratio (AOR) at 95%.

### Ethical Consideration

An official ethical letter was obtained from the SPHMMC, the school of nursing as a whole, as well as from the Addis Ababa health bureau, and was submitted to the selected hospitals. Permission letters were obtained from those bodies prior to data collection. The study participants were informed about the objective and rationale of the study and were guaranteed their choice of participation or refusal at any point in the data collection. And all information was recorded anonymously, and confidentiality was also assured.

## Results

### Socio-demography characteristics of participant

In this study, 414 participants from the selected government hospital participated in the study and were requested for interviews as they fulfilled the inclusion criteria; among those, only 390 (94.2%) completed the interview. The mean age of participants was 56.31 ± 14.61, at which the majority of the respondents, 187 (47.9%), were above 60 years old, and 205 (52.6%) of the participants were female. The majority 317 (81.3%) participants live in urban areas, and 248 (63.6%) are living with their partner. 251 (64.4%) had attained formal education, and 167 (42.8%) get more than 2500 birr per month.

**Table 1.** Socio-demography characteristics of hypertensive patients at public hospitals in Addis Ababa, Ethiopia, 2023 (N=390)

| Variable | Category | Frequency | Percent |
| --- | --- | --- | --- |
| Sex | Male | 185 | 47.4 |
|  | Female | 205 | 52.6 |
| Age | 18-39 | 60 | 15.4 |
|  | 40-59 | 143 | 36.7 |
|  | above 60 | 187 | 47.9 |
| Marital status | Single | 42 | 10.8 |
|  | Married | 248 | 63.6 |
|  | Divorced | 40 | 10.3 |
|  | Widowed | 60 | 15.4 |
| Residence | Rural | 73 | 18.7 |
|  | Urban | 317 | 81.3 |
| Education | can't read and write | 69 | 17.7 |
|  | can read and write | 70 | 17.9 |
|  | Primary | 88 | 22.6 |
|  | Secondary | 85 | 21.8 |
|  | college/university and above | 78 | 20.0 |
| Income per month | less than 1500 | 153 | 39.2 |
|  | 1500-2499 | 70 | 17.9 |
|  | above 2500 | 167 | 42.8 |
|  | Total | 390 | 100 |

### Patient related factors

Among 390 respondents, 222 (56.9%) were treated for greater than five years, and 144 (36.9%) were taking two or more antihypertensive medications per day and from patients who were taking single pills per day 121(31%) were took enalapril. 119 (30.5%) had practiced self-blood pressure monitoring, and among those, most of them (39/10%) measured their blood pressure once a week. And 209 (53.6%) of respondents had a controlled blood pressure status. Regarding comorbidity, almost half (51.8%) of patients had at least one comorbidity; among those 84(21.54%) had DM, while 146 (37.3%) had no comorbidity and 51% had good knowledge about hypertension and its management.

**Table 2.** Hypertensive patients characteristics at public hospitals in Addis Ababa, Ethiopia, 2023 (N=390)

| Variable | Category | Frequency | Percent |
| --- | --- | --- | --- |
| BP status | Uncontrolled | 181 | 46.4 |
|  | Controlled | 209 | 53.6 |
| duration in year since start of treatment | less than 5 | 168 | 43.1 |
|  | greater than 5 | 222 | 56.9 |
| number of pills per day | One | 246 | 63.1 |
|  | 2 and above | 144 | 36.9 |
| comorbidity | no comorbidity | 146 | 37.4 |
|  | one | 202 | 51.8 |
|  | 2 and above | 42 | 10.8 |
| practice of SBPM | No | 271 | 69.5 |
|  | Yes | 119 | 30.5 |
| SBPM frequency | don't measure | 271 | 69.5 |
|  | ≥ One daily | 17 | 4.4 |
|  | Once a week | 39 | 10.0 |
|  | Twice a week | 38 | 9.7 |
|  | Irregularly | 25 | 6.4 |
| Knowledge on hypertension and its management | Poor | 191 | 49.00 |
|  | Good | 199 | 51.00 |
|  | Total | 390 | 100 |

### Organizational related characteristics

Majority of the study participants (329 (84.4%)) used health insurance service and received their medication for free from the hospitals; however, about 163, or 41.8%, of the patients said that the hospitals didn’t have their prescribed medications. And 230(59%) had good communication with their health care provider.

**Table 3.** Hypertension patients’ organizational characteristics at public hospitals in Addis Ababa, Ethiopia, 2023 (N=390)

| Variable | Category | Frequency | Percent |
| --- | --- | --- | --- |
| Communication between patient and health care provider | Poor | 160 | 41.0 |
|  | Good | 230 | 59.0 |
|  | Total | 390 | 100 |
| Insurance service user | No | 61 | 15.6 |
|  | Yes | 329 | 84.4 |
|  | Total | 390 | 100 |
| anti-hypertension drug availability in the hospital | No | 163 | 41.8 |
|  | Yes | 227 | 58.2 |
|  | Total | 390 | 100 |

### Respondents’ characteristics on the MMAS-8 scale of medication adherence

To determine the adherence status with MMAS-8; one to seven items were rated as "yes" = 1 and "no" = 0, with the exception of the 5th question, which was changed to "no" = 1, "yes" = 0. On the eighth item, respondents who chose "Never"=1, "Once"=0.25, "Sometimes"=0.75, "Usually"=0.75, and "Always"=0 were rated. And a score of 6 or higher was considered adherent, while a score of less than 6 was considered non-adherent. And the overall adherence rate to anti-hypertension medication was found to be 66.2%. Out of these participants’, more than half (58.2%) hadn’t forgotten taking their medication in the last two weeks prior to the interview, and about 46.4% hadn’t ever faced remembering to take their medications, and almost all (97.7%) had never stopped taking their ordered pills without a doctor’s order. As well, 97.2% responded that they continued taking their medication even when they felt their blood pressure was under control.

**Table 4.** Hypertensive respondents’ characteristics on the MMAS-8 scale of medication adherence at public hospitals in Addis Ababa, Ethiopia, 2023 (N=390)

| Variable | Response | Frequency | Percent |
| --- | --- | --- | --- |
| Do you ever forget to take your medicine? | Yes | 163 | 41.8 |
|  | No | 227 | 58.2 |
| In the last two weeks, is there any day when you did not take your high blood pressure medication? | Yes | 24 | 6.2 |
|  | No | 366 | 93.8 |
| Have you ever stopped taking your medications or decreased the dose without your doctor order, because you felt worse when you took them? | Yes | 9 | 2.3 |
|  | No | 381 | 97.7 |
| Do you forget to take your medications, when you travel or leave the house? | Yes | 107 | 27.4 |
|  | No | 283 | 72.6 |
| Did you take your high blood pressure medication yesterday? | Yes | 360 | 92.3 |
|  | No | 30 | 7.7 |
| Do you stop taking your medications, when you feel your blood pressure is controlled? | Yes | 11 | 2.8 |
|  | No | 379 | 97.2 |
| Have you ever felt distressed for strictly following your high blood pressure treatment? | Yes | 88 | 22.6 |
|  | No | 302 | 77.4 |
| How often do you have difficulty to remembering taking all you blood pressure medication? | Never | 181 | 46.4 |
|  | Once | 114 | 29.2 |
|  | Sometimes | 90 | 23.1 |
|  | Usually | 4 | 1.0 |
|  | Always | 1 | 0.3 |
| Total | Adhered | 258 | 66.2% |
|  | No-adhered | 132 | 33.8% |

**Figure 1.**
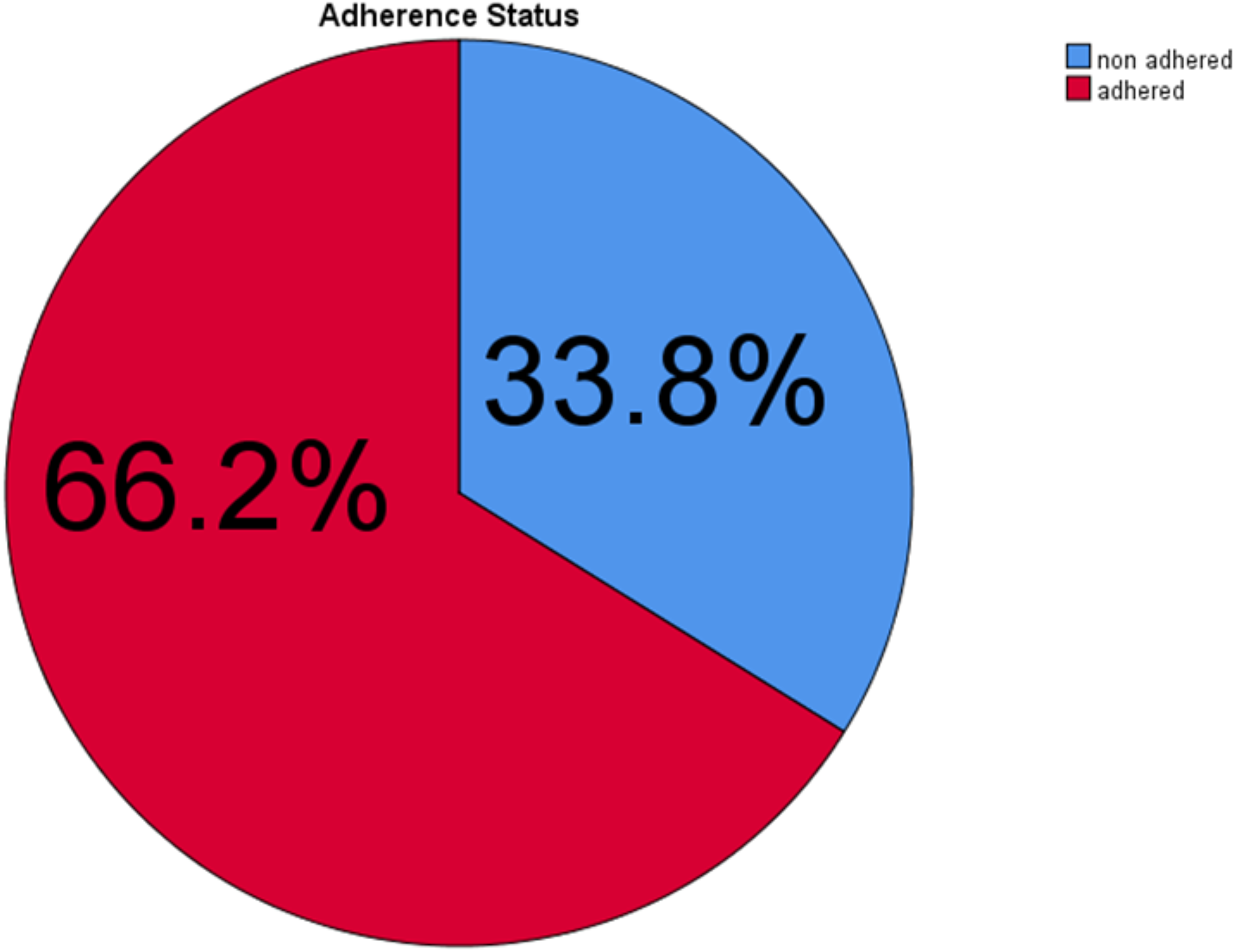
Ant hypertension medication adherence status at Addis Ababa, 2023, Ethiopia (N=390)

### Factors Associated with Antihypertensive Medication Adherence

Using both the bivariate and multivariate logistic regression techniques, the impact of socio-demographic, patient-related factors, and organizational factors on adherence status was analyzed (table 5). Consequently, each of the following independent variables was taken into consideration as a candidate for multivariate analysis after having a p-value less than 0.25: residence; educational status; income; knowledge of HTN and its management; practice of self-blood pressure monitoring; frequency of self-blood pressure monitoring; communication between patients and healthcare providers; use of health insurance services; availability of drugs in the hospital; and BP status.

**Table 5.**
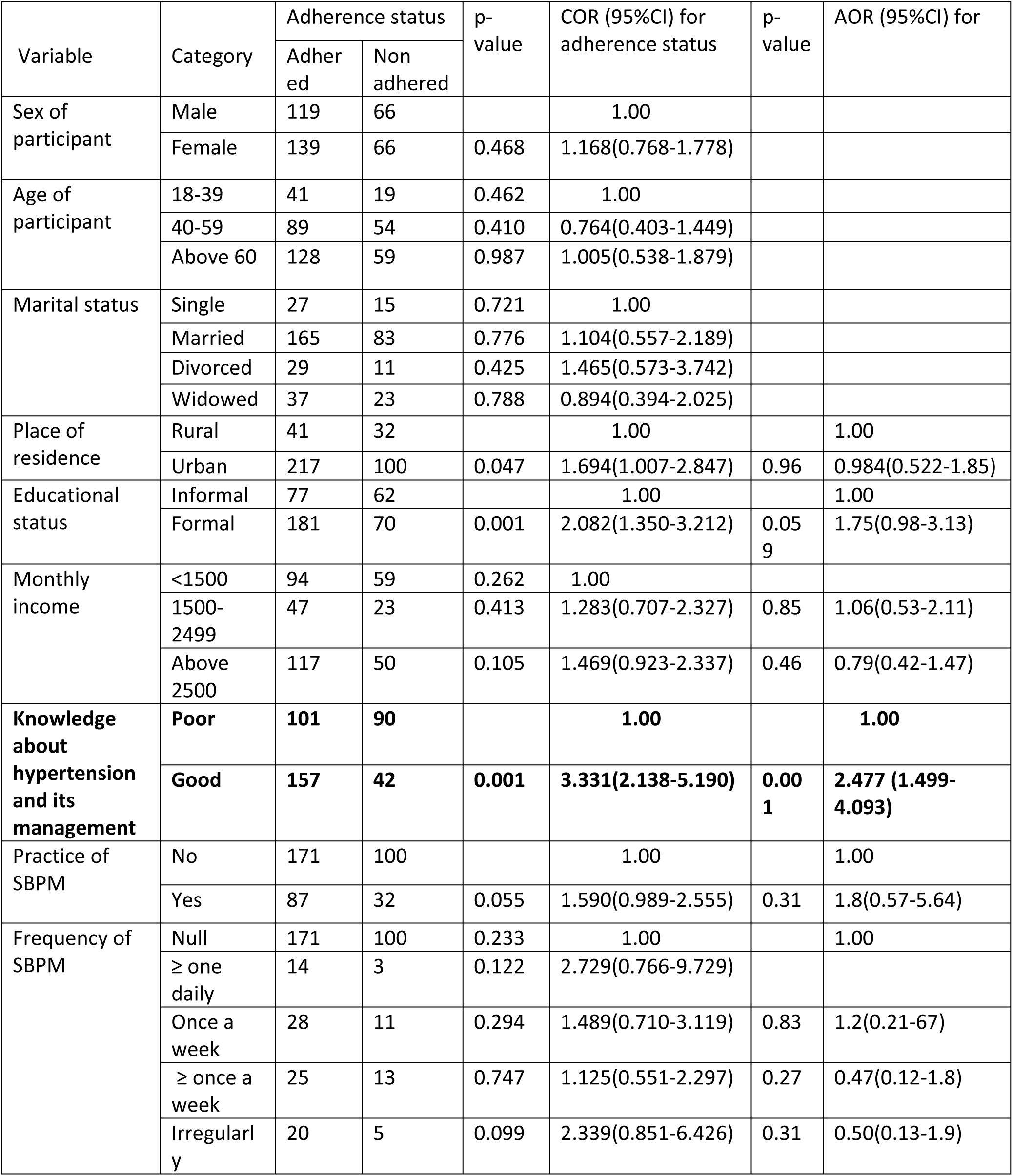
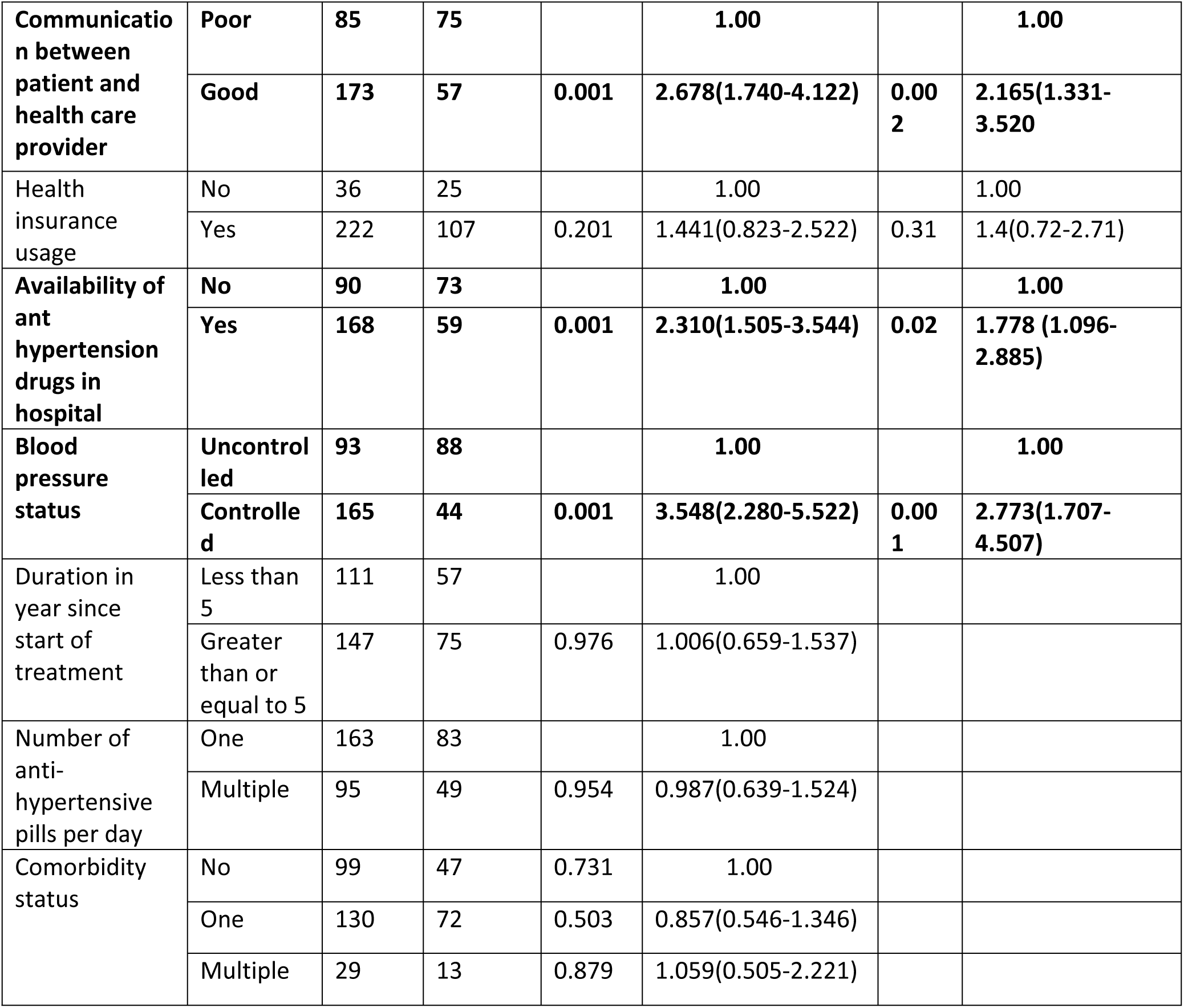
Bivariate and multivariate logistic regression analysis result showing factors associated with ant hypertension medication adherence among hypertensive patients in Addis Ababa, Ethiopia, 2023.

Hosmer-Lemeshow goodness of fit (P = 0.209) and multicollinearity (VIF = 1.08–6) tests were done prior to multivariable analysis, and both proved that the model was well fitted and that there was no multicollinearity among the variables.

And finally patients with good knowledge about HTN and its management, good patient-to-health care provider communication, availability of drugs in the hospital, and controlled BP status remained significantly associated with adherence to ant hypertension medications, with a minimum S.E. of 0.248 and a maximum of 0.256.

So in this study as of (table 5) with the overall adherence rate of 66.2%, respondents who had good knowledge about hypertension and its management was about two times (AOR=2.477, 95%CI (1.499-4.093)) more likely to be adherent than who had poor knowledge. Those who have good communication with their health care provider had a significantly higher chance of being adherent to their medication (AOR=2.165, 95%CI (1.331-3.520)) than those who had poor communication. Similarly those who got ant-hypertension drugs easily in their hospital had higher chance of being adherent (AOR=1.778, 95%CI (1.096-2.885)) than those who didn’t get easily. Patients who had controlled blood pressure were 2.77 times (AOR=2.773, 95% CI (1.707-4.507)) more likely to be adherent than those with uncontrolled blood pressure status.

## Discussion

With the intent of avoiding consequences, patients’ adherence to their antihypertensive drugs is crucial. In many developing countries, ensuring patient adherence to anti-hypertension drugs in order to prevent hypertension-related consequences continues to be a significant problem(5). And this study found that 258 respondents, or about 66.2%, adhered to their anti-hypertension medication treatment. And good knowledge about hypertension and its management, good communication with health care providers, easy access to ant hypertension drugs, and having controlled blood pressure had a significant association with treatment adherence.

Only 66.2% of the study participants were found to be adherent to their therapy; this percentage is also nearly identical to that of studies conducted in Hawasa referral hospital (67%)(23). The determined adherence status prevalence of this study is also almost as of the previous study conducted in 2016 in Addis Ababa(24) which indicate that no interventions had not been taken or may be due to low improvement of health care quality compared to increased number of patients on follow up.

It is greater than what was reported from Sudan (58.4%)(25), Nedjo General Hospital(31%)(26), and Goba Referral Hospital(24%)(27). The current study’s findings on antihypertensive treatment adherence were also better than the Iran’s(50.7%)(28). This discrepancy may be due to different methods of measurement, study population as well as smaller sample sizes were used in Sudan (202), in Nedjo(171) and Goba referral hospitals(260) in comparison to this study but it falls below of research conducted in Tanzania (76.9%),and South America (75%)(29,30) and this might be because patients have better access to and care in those countries. The study’s findings further support the fact that 41.8% of the participants had difficulty getting their anti-hypertension medications from their hospitals.

People with hypertension who had a basic understanding of HTN and how to control it were two times more likely to maintain compliance with their medication regimen than individuals with poor knowledge and this is in line to study conducted in Kenya(31), as well as in Hareri and Dire Dawa in which knowledgeable patient had about two times more adhered than who had poor knowledge of hypertension(32) while in Hawasa had adhered three times more(23). The potential cause could be that informed patients are becoming more conscious of the consequences of improper drug use.

In this study, it was discovered that patient-provider communication was important for describing adherence to medications. This finding fits in with earlier research from USA and Vietnam countries(13,33) particularly those involving older persons with HTN which found that improved patient-health care provider communication were linked to better quality care and medication adherence. Similar findings have been made in Indonesia and South Korea(15,34). This is due to the fact that effective patient-provider communication during treatment can increase patients’ understanding of HTN and its management, improves their trust and satisfaction with healthcare services, and helps them build their confidence in carrying out recommended behaviors, which improves medication adherence.

This study revealed that patients who get their ant hypertension drugs from their hospitals were more significantly adhered than those who don’t always get prescribed medications from the hospital. And it is comparable to the study reported from Kenya and Tanzania (30,31) it is also similar as of eastern Ethiopian’s report(32). This could be a result of the easy accessibility of the prescribed drugs during the follow-up period. As well, the majorities (84.4%) of respondents were health insurance users and got drugs for free.

Likewise among 390 patients, those with controlled blood pressure, 53.6%, were 2.77 times more significantly associated with adherence than those with uncontrolled blood pressure, and this finding is nearly identical to that from Pakistan, where respondents with controlled blood pressure adhered to their medication at about three times higher than those with uncontrolled blood pressure(6). Similar situations occurred in Tanzania(30) as well as in Ethiopia(4,23). This may be as a result of correct usage of prescribed antihypertensive drugs.

## Limitation of the study

As mentioned earlier, this study was conducted only in Governmental hospital which was not considering the private, the other is the study was done cross sectionally for one month. So, patients coming for treatment of hypertension may not be included in the study after one month of the study period.

## Conclusion

The study’s adherence prevalence was sub-optimal, and a good patient-to-health care provider communications was significantly related, it has the power to alter medication use. So the design of a patient-to-provider dialogue is therefore crucial for the successful medication adherence of hypertension patients because these patients may want satisfaction from clinicians. Additionally, there was a significant association between having controlled blood pressure, knowing how to manage hypertension, and having a supply of anti-hypertension medication in the hospitals and medication adherence.

## Recommendations

Doctors, nurses, pharmacists, and other healthcare providers work at public hospitals in Addis Ababa must enhance patient interaction in order to give patients more effective treatment and give them the chance to be involved in decision-making and share their opinions about their disease and its management.

As well as health care service administrators found at public hospitals in Addis Ababa also must develop better procedures for dealing with patients, clinicians and other staff members in order to improve patients’ compliance with their medications.

Researchers who are interested on factors of medication adherence also suggested to study on proper use and practice of self-blood pressure measurement among hypertensive patients.

## Data Availability

The data underlying to this research work result is available within the authors and will be immediately available up on request.

## Acknowledgments

We would like to express my deepest heartfelt gratitude to St. Paul hospital millennium medical college, School of nursing for providing the opportunity. We need to express our heartfelt indebt gratitude to Ayalnesh Zemene (MSc, Assistant Professor, Ph.D.) and Habtamu Ayalew (BSc, MSc), Habtamu Mulu (BSc, MSc) for their valuable advice and continued encouragement throughout the development of this research thesis. Lastly, we would like to acknowledge the selected hospitals, health care providers in hospitals for their willingness to provide valuable data.

## Declaration

### Ethics approval and consent to participate

The study was approved by the ethical committee of Saint Paul’s hospital millennium medical college or institutional review board (IRB) of Saint Paul’s hospital millennium medical college with the reference number PM 23/448 at the date of 21/2/23 as well as the informed consent was obtained from Ras desta Damtew Hospital, Yekatit 12 Medical College Hospital written with the reference number of አ/አ/ጤ/1036/227 at the date of march 13/2023, st.peter specialized hospital and st.paulose hospital millennium medical college written with the reference number of N/S/50460 at the date of February 27/2023.

## Abbreviations

AOR: Adjusted Odd Ratio: Blood Pressure
CI: Confidence Interval
CVD: Cardiovascular disease
HTN: Hypertension
MMAS-8: Eight items Morisky’s Medication Adherence Scale
MmHg: millimeter of mercury
SBPM: Self-blood pressure monitoring
SPSS: Statistical Package for Social Science
SPHMMC: St. Paul hospital millennium medical college
SRS: Systematic Random Sampling
SSA: Sub Saharan African
WHO: World health organization

## Data Sharing Statement

The datasets used during this study are available from the principal investigator upon reasonable request.

## Author Contributions

**Habtamu Ayalew:** was involved in proposal writing designed the study and participated in coordination supervised and the overall implementation of the project analyzed data draft and finalized the manuscript.

**Ayalnesh Zemene:** Participated in designing the study supervision and critical revision of the manuscript.

**Habtamu Mulu:** Contributed in data collection and cleaning and analyzing the data

All read and approved the final manuscript gave final approval of the version to be published; agreed on the journal to which the article was submitted; and agree to be accountable for all aspects of the work.

